# Artificial intelligence-driven ECG biomarkers for screening of large pericardial effusion

**DOI:** 10.64898/2025.12.22.25342872

**Authors:** Moon-Seung Soh, Yangyoun Lee, GaYoung Kim, MokEun Park, Hong-Seok Lim, Joon-Han Shin, Youngjin Cho, Joonghee Kim

**Author notes:** **Correspondence:** Professor Joonghee Kim, MD, PhD, Department of Emergency Medicine, Seoul National University Bundang Hospital, 82, Gumi-ro 173 Beon-gil, Bundang-gu, Seongnam, Gyeonggi-do, 13620, Republic of Korea.

## Abstract

**Background:** Pericardial effusion can progress to life-threatening cardiac tamponade when large or rapidly accumulating, yet early diagnosis is frequently delayed without sufficient clinical vigilance. Echocardiography is the gold standard but is not always accessible in emergency or resource-limited settings. Electrocardiographic (ECG) findings, including low QRS voltage and electrical alternans, may prompt clinical suspicion; however, their diagnostic value remains constrained by poor quantitative standardization and limited objectivity.

**Objective:** This study aimed to evaluate the diagnostic performance of *ECG Buddy™*, a smartphone-based artificial intelligence (AI) ECG analysis application, in detecting large pericardial effusion (LPE) and cardiac tamponade.

**Methods:** We retrospectively reviewed adult patients (≥19 years) who underwent echocardiography between January 2013 and December 2023. LPE was defined as an effusion ≥2.0 cm in maximal diameter, and tamponade was defined as LPE with constrictive physiology or right atrium/right ventricle (RA/RV) diastolic collapse. An equal number of control patients without LPE were randomly sampled. Printed ECGs were analyzed using *ECG Buddy™*, which generated a quantitative digital biomarker (QCG-LPE). Diagnostic performance was evaluated with receiver operating characteristic-area under the curve (ROC-AUC), sensitivity, specificity, and comparison with conventional biomarkers (Troponin I, NT-proBNP). Biomarker changes before and after pericardiocentesis (PCC) were also assessed.

**Results:** A total of 770 patients (LPE=385, controls=385) were included, of whom 112 had tamponade and 158 underwent PCC. QCG-LPE levels differed markedly across groups: Normal 1.0 (IQR 0.3–3.5), LPE 37.8 (21.2–62.1), and Tamponade 61.8 (38.3–80.0) (p<0.001). QCG-LPE achieved an AUC of 0.980(95% CI, 0.972–0.988) for detecting LPE, with sensitivity 96.6% (95% CI, 94.3–98.7) and specificity 88.4% (95% CI, 84.9–91.6). For tamponade, the AUC was 0.880 (95% CI, 0.851–0.909), sensitivity 94.0% (95% CI, 88.1–98.8) and specificity 73.4% (95% CI, 69.7–77.0). QCG-LPE outperformed both Troponin I (AUC 0.731) and NT-proBNP (AUC 0.920) with a statistically significant difference (p<0.001). In patients undergoing PCC, biomarker values decreased significantly after the procedure (48.95 → 22.95, p<0.001), demonstrating responsiveness to therapeutic intervention.

**Conclusions:** Digital ECG biomarker demonstrated excellent diagnostic accuracy for LPE and tamponade, validated against echocardiography as the gold standard. Its quantitative biomarker not only enables rapid screening but also reflects therapeutic response, suggesting potential for use as an accessible adjunct tool in emergency and resource-limited settings.

## Introduction

Pericardial effusion refers to the accumulation of fluid within the pericardial sac. While small to moderate effusions may remain clinically silent, a large amount of effusion or a rapidly accumulating effusion can increase intrapericardial pressure and compromise cardiac filling, ultimately leading to **cardiac tamponade**.[1,2] Cardiac tamponade is a medical emergency characterized by impaired ventricular filling, reduced cardiac output, and, if not promptly recognized and treated, hemodynamic collapse and mortality. Because of its potentially catastrophic consequences, timely detection of clinically significant pericardial effusion is essential.

The diagnosis of tamponade is often challenging. Symptoms and signs such as dyspnea, hypotension, tachycardia, and jugular venous distension are nonspecific and can overlap with other acute cardiovascular or pulmonary conditions. Echocardiography is the **gold standard** for evaluating pericardial effusion and identifying tamponade physiology, providing detailed information of effusion size, right-sided chamber collapse, and hemodynamic significance.[3–5] However, echocardiography requires equipment and skilled personnel, which may not always be available in emergency departments or in resource-limited settings. This limitation underscores a critical demand for accessible, rapid, and quantitatively robust diagnostic tools to facilitate timely clinical decision-making.

Electrocardiography (ECG) is routinely performed as a first-line test in patients presenting with dyspnea, chest pain, or hemodynamic instability. Classical ECG findings associated with tamponade include **low QRS voltage** and **electrical alternans**.[1,6] Although these signs may suggest the presence of a large effusion or tamponade, they are qualitative in nature, lack sensitivity and specificity, and cannot provide an objective or standardized measure of risk.[7,8] As a result, tamponade may be underrecognized or misdiagnosed when echocardiography is not readily available.

Recent advances in artificial intelligence (AI) have enabled novel applications of ECG analysis, particularly through deep learning models trained on large datasets. AI-enhanced ECG interpretation has shown promise in detecting conditions such as left and right ventricular dysfunction, myocardial infarction, and hyperkalemia using either digital waveform data or printed ECG images.[9–11] In this context, a smartphone-based AI application, *ECG Buddy™*, has been developed and validated for analyzing printed 12-lead ECGs by converting them into quantitative ECG biomarkers (QCG™ scores).[12–22] This technology bypasses the need for direct integration with ECG machines or electronic medical records, making it more adaptable to real-world clinical environments where only printed or displayed ECGs are available.

Given the clinical importance of rapidly identifying patients at risk of tamponade in the limitations of both traditional ECG interpretation and echocardiography availability, the evaluation of *ECG Buddy™* for the detection of large pericardial effusion (LPE) and tamponade is of substantial clinical relevance. In this study, we aimed to validate the performance of a digital ECG biomarker derived from *ECG Buddy™* in identifying LPE and cardiac tamponade.

## Methods

### Study Design

This retrospective study was conducted at a tertiary academic hospital and included adult patients (≥19 years) who underwent transthoracic echocardiography between January 2013 and December 2023. The institutional review board approved the study protocol and waived the requirement for informed consent due to the retrospective nature of the study (AJOUIRB-SW-2024-488).

### Study Goal

The primary goal of this study was to evaluate the diagnostic accuracy of the QCG-LPE biomarker generated by ECG Buddy™ for detecting LPE or cardiac tamponade, with echocardiography serving as the gold standard reference. Secondary goals were: (1) to assess the diagnostic accuracy of QCG-LPE for distinguishing cardiac tamponade from mixed population of LPE and non-LPE, (2) to compare its performance with conventional laboratory biomarkers including Troponin I and NT-proBNP, and (3) to examine changes in QCG-LPE before and after pericardiocentesis (PCC).

### Eligibility Criteria

Patients were eligible if echocardiography demonstrated either (1) LPE or (2) cardiac tamponade. Large pericardial effusion was defined as a maximal effusion diameter ≥2.0 cm on echocardiography. Cardiac tamponade was defined as LPE accompanied by at least one of the following: (a) right atrial or right ventricular diastolic collapse, or (b) constrictive physiology such as inferior vena cava dilation >2.0 cm with <50% inspiratory collapse, or >30% respiratory variation in mitral inflow. For the control group, patients who showed no evidence of LPE or cardiac tamponade on echocardiography during the same study period were selected.

### AI Analysis

In this study, ECG images were analyzed with ECG Buddy™. It is a smartphone-based AI software approved by the Korean Ministry of Food and Drug Safety (MFDS) as a Class II medical device.[23] The application analyzes printed 12-lead ECG images and produces quantitative ECG biomarkers (QCG™ score) ranging from 0 to 100. These biomarkers are correlated with the risk of specific emergencies and cardiac dysfunctions, including acute coronary syndrome, hyperkalemia, and LPE or cardiac tamponade. In patients who underwent PCC, only the pre-procedure biomarker was used for diagnostic performance evaluation, while paired pre- and post-procedure ECGs were used to assess biomarker response.

### Statistical Analysis

Continuous variables were reported as medians with interquartile ranges (IQR) and categorical variables as counts with percentages. Group comparisons were performed using the Mann–Whitney U test or chi-square test, as appropriate. Diagnostic performance was evaluated using **receiver operating characteristic (ROC) curve analysis**, with area under the curve (AUC) and 95% confidence intervals (CI). Optimal cut-off thresholds were determined by the **Youden index**, and sensitivity, specificity, and odds ratio (OR) were calculated. AUCs between biomarkers were compared using **DeLong’s test**. For patients undergoing PCC, paired comparisons of biomarker levels before and after the procedure were made using the **Wilcoxon signed-rank test**. Additionally, **subgroup analyses** were performed according to **sex (male vs female)** and **age (**≥**65 years vs <65 years)**. All statistical tests were two-tailed, and a p-value <0.05 was considered statistically significant. All analyses were conducted with R software (version 4.1.0; R Foundation for Statistical Computing, Vienna, Austria).

## Results

### Patient Characteristics

During the study period, a total of **770 patients** were included (LPE=385, controls=385). Among them, **112 patients (29.1%)** had cardiac tamponade, and **158 patients (41.0%)** underwent PCC. Baseline characteristics are summarized in **Table 1**. Patients with LPE were older (median 68.0 vs. 59.0 years, p<0.001) and showed higher diastolic blood pressure (78.0 vs 75.0, p=0.004) and heart rate (89.0 vs. 71.0, p<0.001) than control group. They also had higher rates of diabetes mellitus (34.8% vs. 21.6%, p<0.001), hypertension (61.3% vs. 39.2%, p<0.001), cerebrovascular disease (14.3% vs. 2.3%, p<0.001), and end-stage renal disease (17.7% vs. 1.6%, p<0.001). Laboratory findings also revealed significant differences, with higher troponin I and NT-proBNP levels in the LPE group (both p<0.001).

**Table 1.**
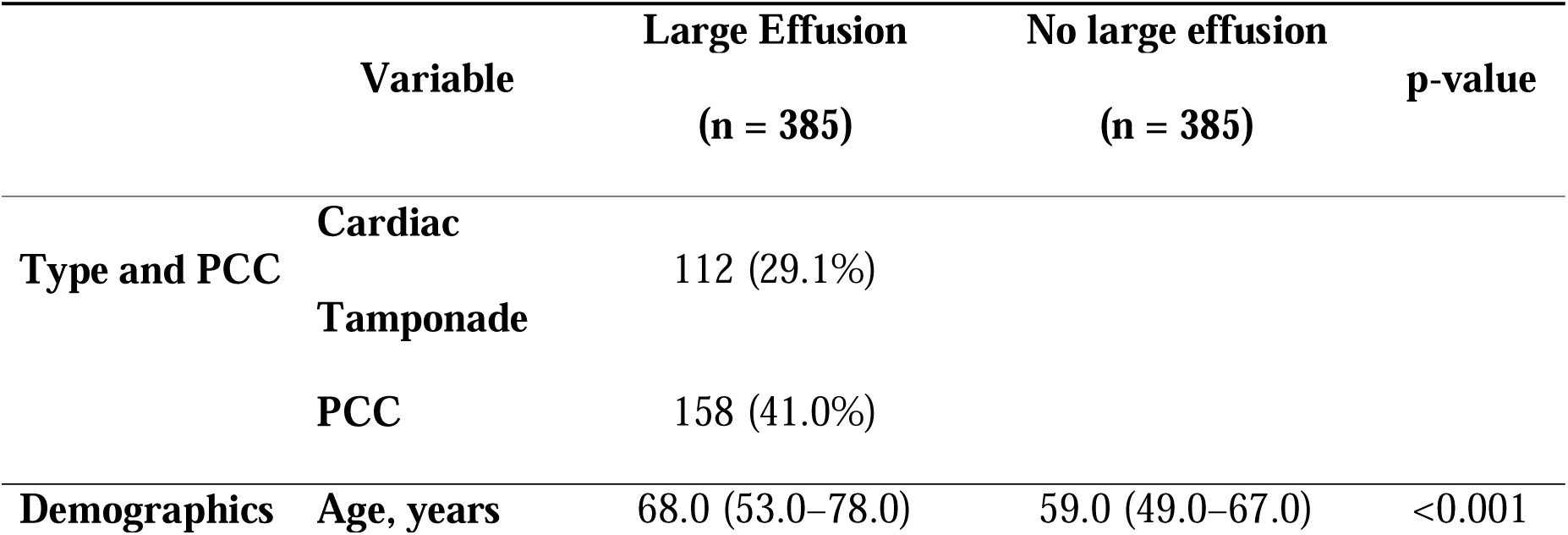
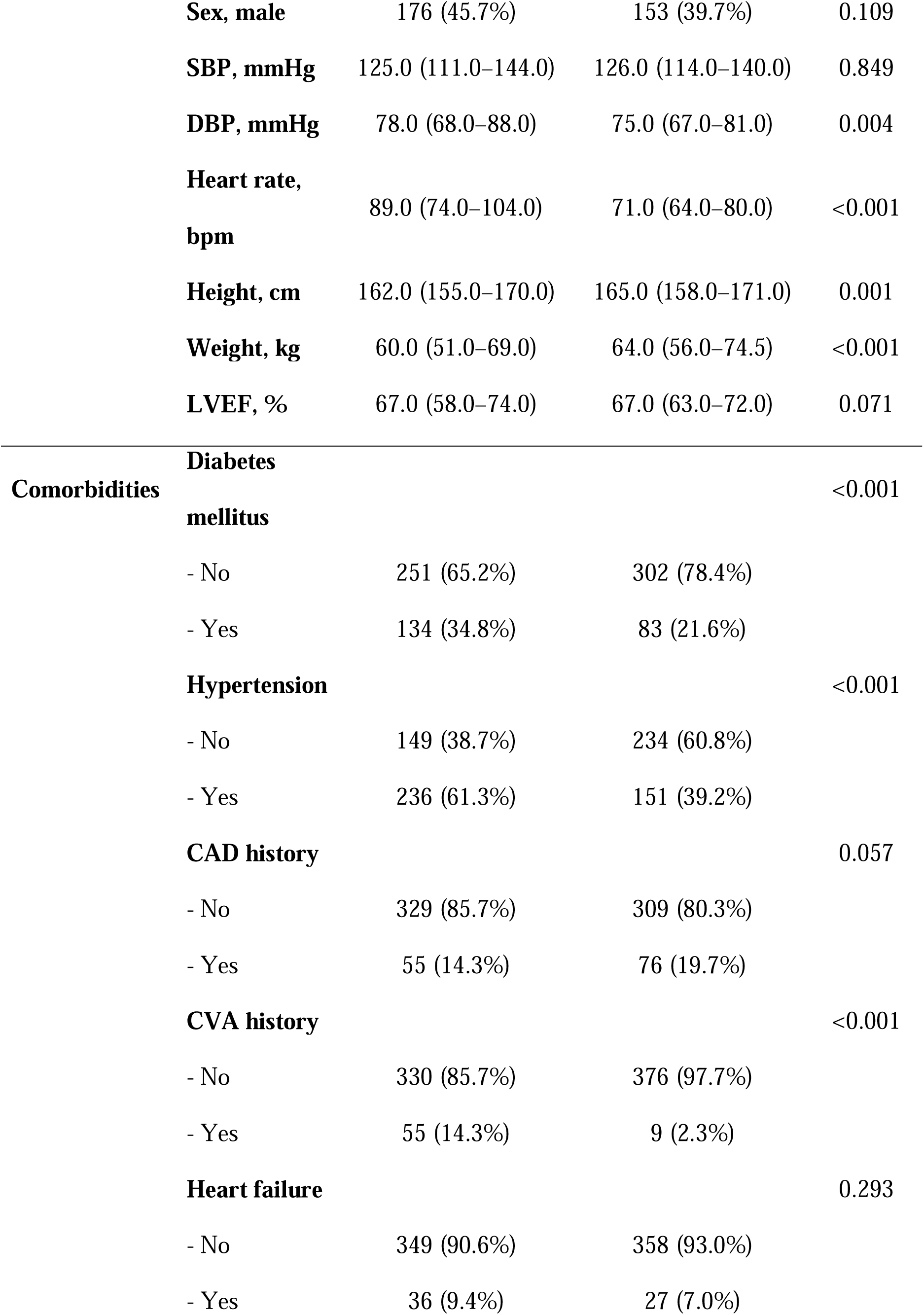
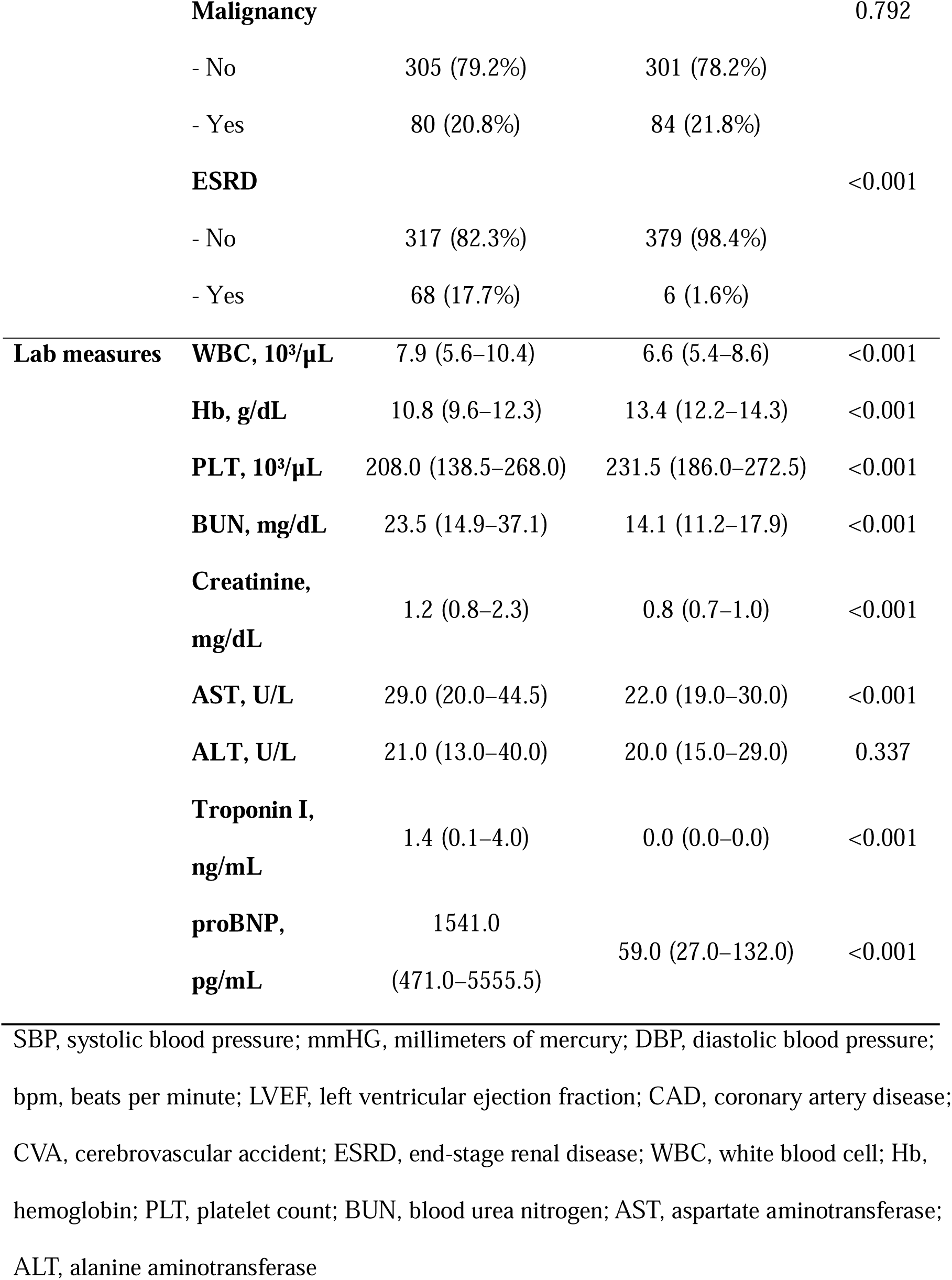
Patient Characteristics.

### QCG™-LPE biomarker Difference

Pre-procedure ECGs were available for **344 controls, 213 LPE patients, and 84 tamponade patients**, which were used for biomarker performance analysis. The QCG™-LPE score differed significantly across groups (p<0.001). Median values were **Normal group: 1.0 (IQR 0.3–3.5)**, **LPE group: 37.8 (IQR 21.2–62.1)**, and **Tamponade group: 61.8 (IQR 38.3–80.0)** (**Figure 1**).

**Figure 1.**
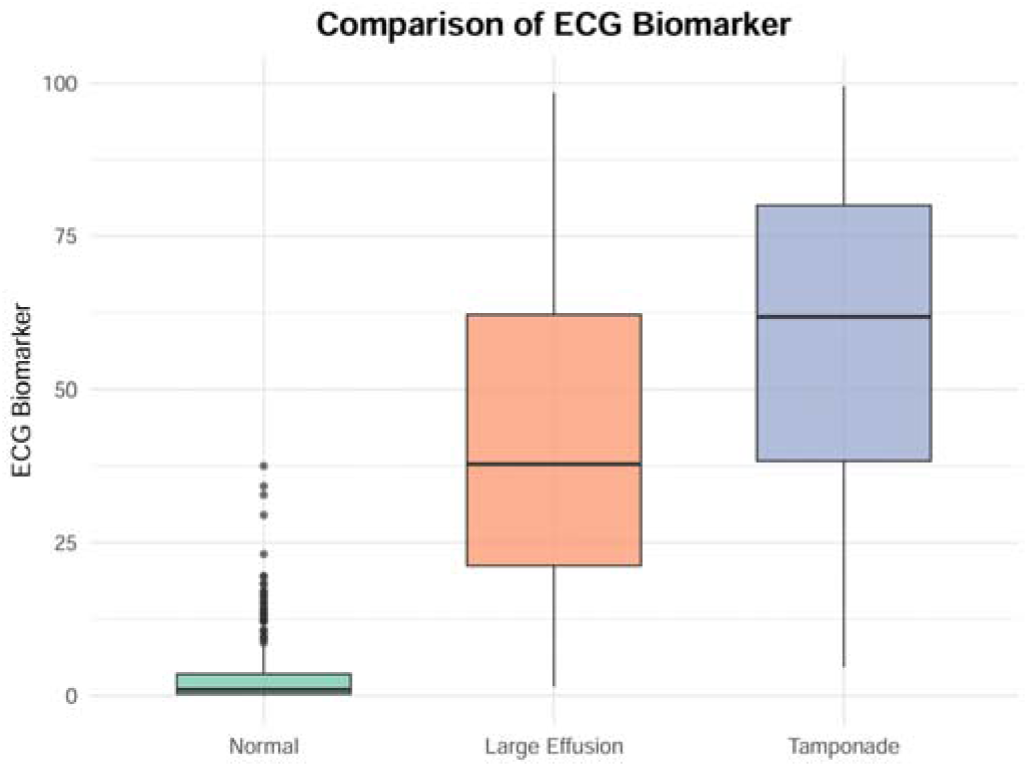
ECG Biomarker Difference. There were significant differences in the level of the pre-procedure digital biomarker: Normal: 1.0 (IQR, 0.3-3.5), Large Effusion: 37.8 (IQR, 21.2-62.1), Cardiac Tamponade: 61.8 (IQR, 38.3-80.0) with P for difference <0.001.

For detecting LPE, QCG-LPE achieved an **AUC of 0.980 (95% CI, 0.972–0.988)**, with **sensitivity 96.6% (95% CI, 94.3–98.7)** and **specificity 88.4% (95% CI, 84.9–91.6)** at the optimal cut-off of 7.65 (Table 2, **Figure 2**). For detecting cardiac tamponade, QCG-LPE yielded an **AUC of 0.880 (95% CI, 0.851–0.909)**, with **sensitivity 94.0% (95% CI, 88.1–98.8)** and **specificity 73.4% (95% CI, 69.7–77.0)** at higher cut-off (26.75).

**Figure 2.**
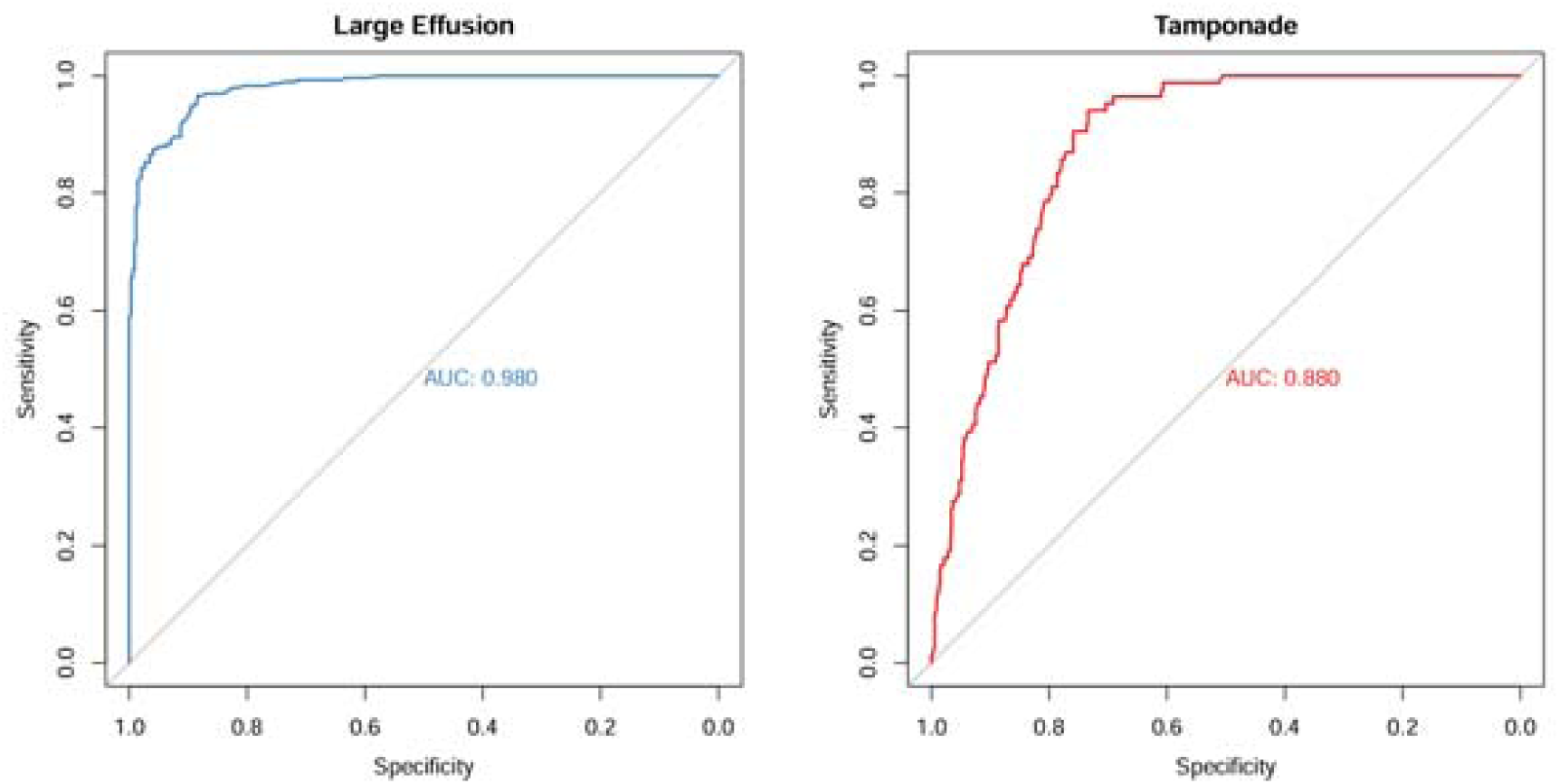
Receiver operating characteristic curve of the ECG Biomarker for Large Pericardial Effusion (Left) and Cardiac Tamponade (Right).

**Table 2.** Diagnostic Performance of ECG Biomarker.

| Outcomes | AUC | Threshold | TP | TN | FP | FN | Sensitivity<br>(95% CI) | Specificity<br>(95% CI) | Odds Ratio<br>(95% CI) |
| --- | --- | --- | --- | --- | --- | --- | --- | --- | --- |
| Large Pericardial<br>Effusion | 0.980<br>(0.972-0.988) | 7.65 | 287 | 304 | 40 | 10 | 96.6<br>(94.3-98.7) | 88.4<br>(84.9-91.6) | 218.12<br>(107.08-444.31) |
| Cardiac<br>Tamponade | 0.880<br>(0.851-0.909) | 26.75 | 79 | 409 | 148 | 5 | 94.0<br>(88.1-98.8) | 73.4<br>(69.7-77.0) | 43.66<br>(17.35-109.91) |

### Comparison with Conventional Biomarkers

When compared with conventional cardiac biomarkers, QCG-LPE outperformed both Troponin I (AUC 0.731, 95% CI 0.677–0.785) **and** NT-proBNP (AUC 0.920, 95% CI **0.889–0.952)** with statistically significant differences (p<0.001 for both comparisons) (Table 3).

**Table 3.** Comparison with Conventional Cardiac Biomarkers.

| Outcomes | AUC | Threshold | TP | TN | FP | FN | Sensitivity<br>(95% CI) | Specificity<br>(95% CI) | Odds Ratio<br>(95% CI) | p-value<sup>1</sup> |
| --- | --- | --- | --- | --- | --- | --- | --- | --- | --- | --- |
| LV Ejection Fraction | 0.564<br>(0.516-0.612) | 57.5 | 86 | 261 | 10 | 207 | 29.4<br>(24.2-34.8) | 96.3<br>(93.7-98.5) | 0.09<br>(0.05-0.18) | <0.001 |
| Troponin I | 0.731<br>(0.677-0.785) | 0.0165 | 177 | 138 | 36 | 45 | 79.7<br>(74.8-85.1) | 79.3<br>(73.0-85.1) | 15.08<br>(9.22-24.65) | <0.001 |
| NT-ProBNP | 0.920<br>(0.889-0.952) | 459 | 118 | 133 | 7 | 38 | 75.6<br>(68.6-82.1) | 95.0<br>(91.4-98.6) | 59.00<br>(25.39-137.13) | <0.001 |

### QCG™-LPE biomarker Response to Pericardiocentesis

Among patients who underwent PCC, the QCG-LPE biomarker demonstrated a marked reduction following the procedure, consistent with both a decrease in effusion volume and concomitant clinical improvement. The median biomarker value declined from 48.95 (IQR, 33.40–73.75) prior to PCC to 22.95 (IQR, 12.75–43.08) after PCC (p < 0.001; **Figure 3**).

**Figure 3.**
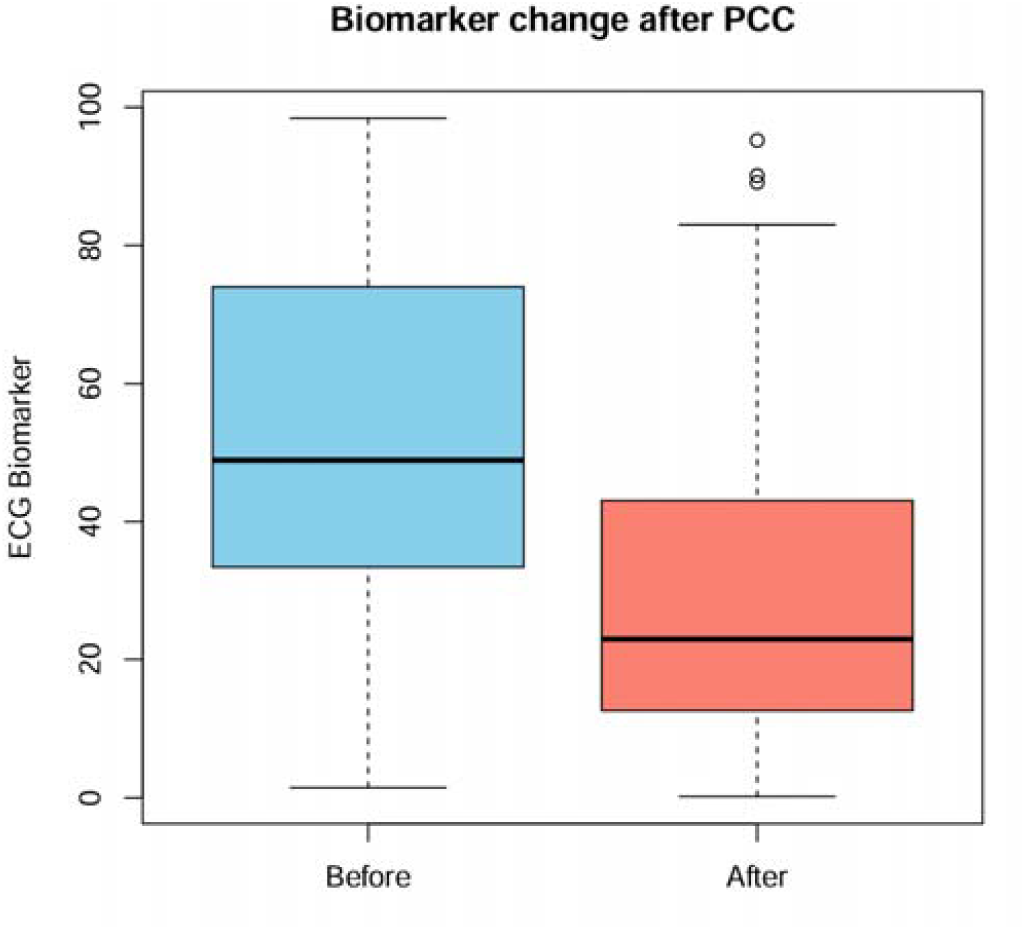
ECG Biomarker Change after PCC. Application of PCC was associated with decrease in the digital biomarker. Biomarker before PCC at 48.95 (IQR, 33.40-73.75) and after at 22.950 (IQR, 12.750-43.075) with P<0.001 (pairwise comparison).

### Subgroup Analysis

Subgroup analyses shown in **Table 4** confirm that QCG-LPE maintains high diagnostic accuracy, exhibiting consistently elevated AUCs and stable performance independent of both sex and age.

**Table 4.** Subgroup Analysis.

| Subgroup | AUC | Threshold | TP | TN | FP | FN | Sensitivity<br>(95% CI) | Specificity<br>(95% CI) | Odds Ratio<br>(95% CI) |
| --- | --- | --- | --- | --- | --- | --- | --- | --- | --- |
| Male | 0.989<br>(0.979-0.998) | 7.65 | 159 | 69 | 4 | 6 | 96.4<br>(93.3-98.8) | 94.5<br>(89.0-98.6) | 457.12<br>(125.04-1671.15) |
| Female | 0.978<br>(0.963-0.993) | 15.5 | 120 | 87 | 4 | 12 | 90.9<br>(85.6-95.5) | 95.6<br>(91.2-98.9) | 217.50<br>(67.86-697.12) |
| Old<br>(≥65) | 0.964<br>(0.941-0.986) | 18.65 | 141 | 55 | 2 | 26 | 84.4<br>(78.4-89.8) | 96.5<br>(91.2-100.0) | 149.13<br>(34.24-649.65) |
| Young<br>(<65) | 0.990<br>(0.982-0.998) | 7.7 | 123 | 102 | 4 | 6 | 95.3<br>(91.5-98.4) | 96.2<br>(92.5-99.1) | 522.75<br>(143.60-1903.01) |

## Discussion

In this retrospective study, we demonstrated that a smartphone-based AI ECG analysis application (*ECG Buddy™*) can accurately identify LPE and cardiac tamponade using printed ECGs. The QCG-LPE biomarker achieved an AUC of 0.980 for LPE detection with sensitivity of 96.6% and specificity of 88.4%, and an AUC of 0.880 for tamponade detection with sensitivity of 94.0% and specificity of 73.4%. The digital biomarker significantly outperformed conventional cardiac marker such as troponin I and NT-proBNP, and its values declined after PCC, reflecting therapeutic response. These findings suggest that ECG-based AI biomarkers may serve as a valuable adjunctive tool for rapid screening and monitoring of patients at risk of tamponade.

### Challenges in Early Tamponade Recognition: Implications of QCG-LPE

Cardiac tamponade represents the most severe manifestation of pericardial effusion physiology, in which elevated pericardial pressure restricts cardiac diastolic filling. Classic clinical findings include hypotension, tachycardia, jugular venous distension, muffled heart sounds (Beck’s triad), and pulsus paradoxus. However, these signs are neither sensitive nor specific, particularly in critically ill patients with multiple comorbidities. Echocardiography remains the most reliable diagnostic tool, identifying features such as right atrial or ventricular diastolic collapse, dilated inferior vena cava with reduced inspiratory collapse, and exaggerated respiratory variation in transvalvular flows.[3,24] Because tamponade can progress rapidly and result in hemodynamic collapse, timely recognition is essential. Yet in clinical practice, diagnosis may be delayed or missed if not suspected early, especially where echocardiography is not immediately available. ECG findings such as low QRS voltage and electrical alternans can raise suspicion, but these qualitative signs lack objectivity and standardized thresholds. In this context, our findings demonstrate that QCG-LPE provides a quantitative and reproducible measure from printed ECGs, enabling standardized risk stratification and supporting clinical decision-making in scenarios where echocardiography is limited.

### Expanding the Clinical Utility of AI-Enabled ECG for Pericardial Disease

Previous studies have shown the potential of AI-enabled ECGs for detecting various critical conditions including ST-elevation myocardial infarction, hyperkalemia and RV strain in pulmonary embolism.[10,11,14,17,25–28] However, few investigations have focused on pericardial effusion and tamponade, particularly using printed ECG images. Our findings expand the applicability of AI ECG analysis by demonstrating excellent diagnostic performance for a structural and hemodynamic disorder beyond conventional arrhythmia or ischemia detection.

A noteworthy finding of our study is that NT-proBNP also showed relatively high diagnostic performance (AUC 0.920) for detecting LPE, likely reflecting the close pathophysiologic link between pericardial constraint, elevated intracardiac filling pressures, and natriuretic peptide release. However, NT-proBNP levels are substantially affected by renal dysfunction, chronic heart failure, and other comorbid conditions, limiting its specificity in real-world settings. In contrast, QCG-LPE demonstrated superior accuracy (p < 0.001) with immediate point-of-care availability, highlighting the practical advantage of AI-based ECG analysis as a more streamlined screening tool.

The clinical utility of this technology may be particularly relevant in emergency departments, prehospital care, or hospitals without 24-hour echocardiography availability. A high QCG-LPE score could prompt urgent echocardiography or early referral, while a low score may help safely exclude significant effusion, streamlining patient triage and resource allocation. In addition, integration into electronic medical records or use by frontline clinicians with limited cardiology expertise may broaden access to timely diagnosis.

### Limitations

This study has several limitations. First, it was a retrospective, single-center analysis, which may limit generalizability. Second, although echocardiography was used as the gold standard, operator variability and report-based classification may introduce bias. Third, only patients with available ECGs were analyzed, which may have introduced selection bias. Finally, while we demonstrated biomarker response to PCC, the prognostic value of QCG-LPE in predicting outcomes such as hemodynamic deterioration or mortality was not assessed.

### Future Directions

Prospective multicenter studies are warranted to validate these findings across diverse populations and healthcare settings. The incorporation of QCG-LPE into real-time clinical workflows, including integration with electronic health records or use in prehospital triage, should be explored. Furthermore, evaluating its role in prognostication and guiding clinical decision-making—such as timing of PCC or escalation of care—represents an important area for future research.

## Conclusion

We externally validated the performance of ECG Buddy™, a smartphone-based AI ECG analysis application, for detecting LPE and cardiac tamponade, using echocardiography as the gold standard. The application showed excellent diagnostic accuracy of digital biomarker for both conditions. Its quantitative signature outperformed conventional cardiac biomarkers and captured treatment response, supporting its utility for screening and monitoring in settings where echocardiography is not readily available.

## Data Availability

All data produced in the present study are available upon reasonable request to the authors

## Acknowledgements

This research was partly supported by a grant from the Medical AI Clinic Program through the National IT Industry Promotion Agency, funded by the Ministry of Science Information Communication and Technology (grant number H0904-24-1002), and the Seoul National University Bundang Hospital Research Fund (grant 13-2021-0018).

## Conflicts of Interest

Joonghee Kim developed the algorithm and founded a start-up company ARPI Inc. He is the CEO of the company. Youngjin Cho works for the company as a research director. Otherwise, there is no conflict of interest for the other authors.

## Abbreviations

ECG: electrocardiogram
AI: artificial intelligence
LPE: large pericardial effusion
RA: right atrium
RV: right ventricle
ROC: receiver operating characteristic
AUC: area under the curve
NT-proBNP: N-terminal pro-B-type natriuretic peptide
PCC: pericardiocentesis
IQR: interquartile ranges
MFDS: ministry of food and drug safety
CI: confidence intervals
OR: odds ratio

---

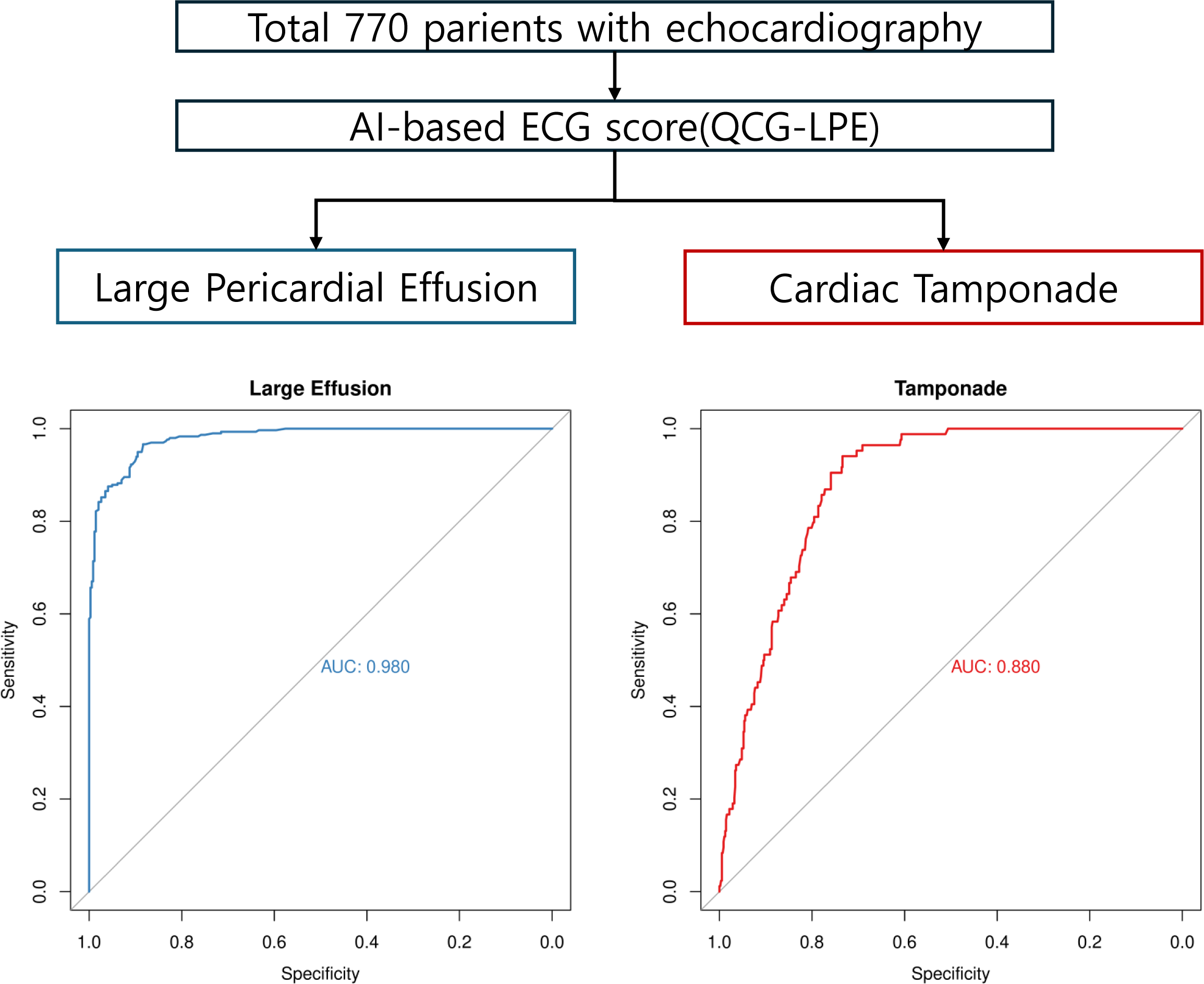

## Notes

### Author Declarations

The institutional review board approved the study protocol and waived the requirement for informed consent due to the retrospective nature of the study (AJOUIRB-SW-2024-488).

## Reference

1. Spodick DH. Acute Cardiac Tamponade. The New England Journal of Medicine. PMID:12917303

2. Reddy PS, Curtiss EI, O’Toole JD, Shaver JA. Cardiac tamponade: hemodynamic observations in man. Circulation 2018;58(2):265–272. PMID:668074

3. Adler Y, Charron P, Imazio M, Badano L, Barón-Esquivias G, Bogaert J, Brucato A, Gueret P, Klingel K, Lionis C, Maisch B, Mayosi B, Pavie A, Ristić AD, Tenas MS, Seferovic P, Swedberg K, Tomkowski W, Achenbach S, Agewall S, Al-Attar N, Ferrer JA, Arad M, Asteggiano R, Bueno H, Caforio ALP, Carerj S, Ceconi C, Evangelista A, Flachskampf F, Giannakoulas G, Gielen S, Habib G, Kolh P, Lambrinou E, Lancellotti P, Lazaros G, Linhart A, Meurin P, Nieman K, Piepoli MF, Price S, Roos-Hesselink J, Roubille F, Ruschitzka F, Sauleda JS, Sousa-Uva M, Voigt JU, Zamorano JL, Zamorano JL, Aboyans V, Achenbach S, Agewall S, Badimon L, Barón-Esquivias G, Baumgartner H, Bax JJ, Bueno H, Carerj S, Dean V, Erol Ç, Fitzimons D, Gaemperli O, Kirchhof P, Kolh P, Lancellotti P, Lip GY, Nihoyannopoulos P, Piepoli MF, Ponikowski P, Roffi M, Torbicki A, Carneiro AV, Windecker S, Shuka N, Sisakian H, Mascherbauer J, Isayev E, Shumavets V, Camp GV, Gatzov P, Hanzevacki JS, Moustra HH, Linhart A, Møller JE, Aboleineen MW, Põder P, Lehtonen J, Antov S, Damy T, Schieffer B, Dimitriadis K, Kiss RG, Rafnsson A, Arad M, Novo S, Mirrakhimov E, Stradinš P, Kavoliuniene A, Codreanu A, Dingli P, Vataman E, Hattaoui ME, Samstad SO, Hoffman P, Lopes LR, Dimulescu DR, Arutyunov GP, Pavlovic M, Dúbrava J, Sauleda JS, Andersson B, Müller H, Bouma BJ, Abaci A, Archbold A, Nesukay E. 2015 ESC Guidelines for the diagnosis and management of pericardial diseases. Eur Hear J 2015;36(42):2921–2964. PMID:26320112

4. Little WC, Freeman GL. Pericardial Disease. Circulation. PMID:16534006

5. Tsang TSM, Enriquez-Sarano M, Freeman WK, Barnes ME, Sinak LJ, Gersh BJ, Bailey KR, Seward JB. Consecutive 1127 Therapeutic Echocardiographically Guided Pericardiocenteses: Clinical Profile, Practice Patterns, and Outcomes Spanning 21 Years. Mayo Clin Proc 2002;77(5):429–436. PMID:12004992

6. Argula RG, Negi SI, Banchs J, Yusuf SW. Role of a 12LLead Electrocardiogram in the Diagnosis of Cardiac Tamponade as Diagnosed by Transthoracic Echocardiography in Patients With Malignant Pericardial Effusion. Clin Cardiol 2015;38(3):139–144. PMID:25694103

7. Eisenberg MJ, Romeral LMD, Heidenreich PA, Schiller NB, Evans GT. The Diagnosis of Pericardial Effusion and Cardiac Tamponade by 12-Lead ECG A Technology Assessment. Chest 1996;110(2):318–324. PMID:8697827

8. Imazio M, Adler Y. Management of pericardial effusion. Eur Hear J 2013;34(16):1186–1197. PMID:23125278

9. Attia ZI, Kapa S, Lopez-Jimenez F, McKie PM, Ladewig DJ, Satam G, Pellikka PA, Enriquez-Sarano M, Noseworthy PA, Munger TM, Asirvatham SJ, Scott CG, Carter RE, Friedman PA. Screening for cardiac contractile dysfunction using an artificial intelligence–enabled electrocardiogram. Nat Med 2019;25(1):70–74. PMID:30617318

10. Al-Zaiti SS, Martin-Gill C, Zègre-Hemsey JK, Bouzid Z, Faramand Z, Alrawashdeh MO, Gregg RE, Helman S, Riek NT, Kraevsky-Phillips K, Clermont G, Akcakaya M, Sereika SM, Dam PV, Smith SW, Birnbaum Y, Saba S, Sejdic E, Callaway CW. Machine learning for ECG diagnosis and risk stratification of occlusion myocardial infarction. Nat Med 2023;29(7):1804–1813. PMID:37386246

11. Galloway CD, Valys AV, Shreibati JB, Treiman DL, Petterson FL, Gundotra VP, Albert DE, Attia ZI, Carter RE, Asirvatham SJ, Ackerman MJ, Noseworthy PA, Dillon JJ, Friedman PA. Development and Validation of a Deep-Learning Model to Screen for Hyperkalemia From the Electrocardiogram. JAMA Cardiol 2019;4(5):428–436. PMID:30942845

12. Kim D, Hwang JE, Cho Y, Cho H-W, Lee W, Lee JH, Oh I-Y, Baek S, Lee E, Kim J. A Retrospective Clinical Evaluation of an Artificial Intelligence Screening Method for Early Detection of STEMI in the Emergency Department. J Korean Méd Sci 2022;37(10):e81. PMID:35289140

13. Choi YJ, Park MJ, Ko Y, Soh M-S, Kim HM, Kim CH, Lee E, Kim J. Artificial intelligence versus physicians on interpretation of printed ECG images: Diagnostic performance of ST-elevation myocardial infarction on electrocardiography. Int J Cardiol 2022;363:6–10. PMID:35691440

14. Kim D, Jeong J, Kim J, Cho Y, Park I, Lee S-M, Oh YT, Baek S, Kang D, Lee E, Jeong B. Hyperkalemia Detection in Emergency Departments Using Initial ECGs: A Smartphone AI ECG Analyzer vs. Board-Certified Physicians. J Korean Méd Sci 2023;38(45):e322. PMID:37987103

15. Kim JH, Jung JY, Kim J, Cho Y, Lee E, Son D. Non-Inferiority Analysis of Electrocardiography Analysis Application vs. Point-of-Care Ultrasound for Screening Left Ventricular Dysfunction. Yonsei Méd J 2024;66(3):172–178. PMID:39999992

16. Park MJ, Choi YJ, Shim M, Cho Y, Park J, Choi J, Kim J, Lee E, Kim S-Y. Performance of ECG-Derived Digital Biomarker for Screening Coronary Occlusion in Resuscitated Out-of-Hospital Cardiac Arrest Patients: A Comparative Study between Artificial Intelligence and a Group of Experts. J Clin Med 2024;13(5):1354. doi: 10.3390/jcm13051354

17. Choi YJ, Park MJ, Cho Y, Kim J, Lee E, Son D, Kim S-Y, Soh MS. Screening for RV Dysfunction Using Smartphone ECG Analysis App: Validation Study with Acute Pulmonary Embolism Patients. J Clin Med 2024;13(16):4792. PMID:39200934

18. Lee SH, Hong WP, Kim J, Cho Y, Lee E. Smartphone AI vs. Medical Experts: A Comparative Study in Prehospital STEMI Diagnosis. Yonsei Méd J 2024;65(3):174–180. PMID:38373837

19. Cho Y, Yoon M, Kim J, Lee JH, Oh I-Y, Lee CJ, Kang S-M, Choi D-J. Artificial Intelligence–Based Electrocardiographic Biomarker for Outcome Prediction in Patients With Acute Heart Failure: Prospective Cohort Study. J Méd Internet Res 2024;26:e52139. PMID:38959500

20. Park J, Kim J, Ahn S, Cho Y, Yoon YE. AI-ECG Supported Decision-Making for Coronary Angiography in Acute Chest Pain: The QCG-AID Study. J Korean Méd Sci 2024;40(12):e105. PMID:40165577

21. Choi J, Kim J, Spaccarotella C, Esposito G, Oh I-Y, Cho Y, Indolfi C. Smartwatch ECG and artificial intelligence in detecting acute coronary syndrome compared to traditional 12-lead ECG. IJC Hear Vasc 2025;56:101573. doi: 10.1016/j.ijcha.2024.101573

22. Lee H, Kwon WY, Song KJ, Jo YH, Kim J, Cho Y, Hwang JE, Choi Y. Interethnic Validation of an ECG Image Analysis Software for Detecting Left Ventricular Dysfunction in Emergency Department Population. Clin Exp Emerg Med 2025; PMID:40304026

23. Ministry of Food and Drug Safety. Available online: https://emedi.mfds.go.kr/search/data/MNU20237 (accessed on 19 December 2025)

24. Klein AL, Abbara S, Agler DA, Appleton CP, Asher CR, Hoit B, Hung J, Garcia MJ, Kronzon I, Oh JK, Rodriguez ER, Schaff HV, Schoenhagen P, Tan CD, White RD. American Society of Echocardiography Clinical Recommendations for Multimodality Cardiovascular Imaging of Patients with Pericardial Disease Endorsed by the Society for Cardiovascular Magnetic Resonance and Society of Cardiovascular Computed Tomography. J Am Soc Echocardiogr 2013;26(9):965–1012.e15. PMID:23998693

25. Liu W-C, Lin C-S, Tsai C-S, Tsao T-P, Cheng C-C, Liou J-T, Lin W-S, Cheng S-M, Lou Y-S, Lee C-C, Lin C. A deep learning algorithm for detecting acute myocardial infarction. EuroIntervention 2021;17(9):765–773. PMID:33840640

26. Wysokinski WE, Meverden RA, Lopez-Jimenez F, Harmon DM, Inojosa BJM, Suarez AB, Liu K, Inojosa JRM, Casanegra AI, McBane RD, Houghton DE. Electrocardiogram Signal Analysis With a Machine Learning Model Predicts the Presence of Pulmonary Embolism With Accuracy Dependent on Embolism Burden. Mayo Clin Proc: Digit Heal 2024;2(3):453–462. PMID:40206108

27. DuBrock HM, Wagner TE, Carlson K, Carpenter CL, Awasthi S, Attia ZI, Frantz RP, Friedman PA, Kapa S, Annis J, Brittain EL, Hemnes AR, Asirvatham SJ, Babu M, Prasad A, Yoo U, Barve R, Selej M, Agron P, Kogan E, Quinn D, Dunnmon P, Khan N, Soundararajan V. An electrocardiogram-based AI algorithm for early detection of pulmonary hypertension. Eur Respir J 2024;64(1):2400192. PMID:38936966

28. Herman R, Meyers HP, Smith SW, Bertolone DT, Leone A, Bermpeis K, Viscusi MM, Belmonte M, Demolder A, Boza V, Vavrik B, Kresnakova V, Iring A, Martonak M, Bahyl J, Kisova T, Schelfaut D, Vanderheyden M, Perl L, Aslanger EK, Hatala R, Wojakowski W, Bartunek J, Barbato E. International evaluation of an artificial intelligence–powered electrocardiogram model detecting acute coronary occlusion myocardial infarction. Eur Hear J - Digit Heal 2023;5(2):123–133. PMID:38505483

